# Plasma cell-free DNA methylomes for hepatocellular carcinoma detection and monitoring after liver resection or transplantation

**DOI:** 10.1101/2024.10.01.24314116

**Authors:** Kui Chen, Zhihao Li, Bianca O. Kirsh, Ping Luo, Stephanie Pedersen, Roxana C. Bucur, Nadia A. Rukavina, Jeffrey P. Bruce, Arnavaz Danesh, Mazdak Riverin, Sandra E. Fischer, Mamatha Bhat, Nazia Selzner, Sonya A. MacParland, Carol-Anne Moulton, Steven Gallinger, Ian D. McGilvray, Mark S. Cattral, Markus Selzner, Trevor W. Reichman, Chaya Shwaartz, Blayne A. Sayed, Sean P. Cleary, Gonzalo Sapisochin, Anand Ghanekar, Trevor J. Pugh

## Abstract

**Background:** Hepatocellular carcinoma (HCC) is one of the most common and lethal malignancies worldwide. HCC diagnosis, monitoring, and treatment decisions rely predominantly on imaging. Curative surgery is limited to those with disease confined to the liver, but recurrence is common. Detection of HCC by mutational profiling of blood plasma cell-free DNA (cfDNA) is limited by mutational heterogeneity and difficulty obtaining tumor tissue to guide targeted gene panels. In contrast, DNA methylation patterns reveal biological processes without need for prior mutational knowledge. We evaluated cell-free methylated DNA immunoprecipitation and high-throughput sequencing (cfMeDIP-Seq) for HCC detection and monitoring of recurrence after curative-intent surgery.

**Methods:** We identified patients undergoing liver transplantation or resection and collected blood at surgery (baseline) and every 3 months for two years (follow-up). We performed cfMeDIP-Seq followed by machine learning to i) develop an HCC classifier based on 300 differentially methylated regions in a Discovery cohort of 35 living liver donors (healthy controls) and 52 baseline samples from HCC patients; ii) test the classifier in a separate Validation cohort of 37 baseline and 112 follow-up samples from 37 patients; and iii) assign an HCC methylation score (HMS) to samples based on their probability (0.0-1.0) of containing HCC-derived cfDNA. We assessed the relationships between HMS and clinical variables.

**Results:** cfMeDIP-Seq to a depth of 101-129 (median 113) million reads per sample succeeded in 201 plasma samples from 89 HCC patients (57 transplant and 32 resection) and 35 healthy controls. In the Discovery cohort, the HCC classifier identified HCC with 97% sensitivity and 99% specificity (mean AUROC = 0.999). In the Validation cohort, the classifier identified HCC with 97% accuracy and HMS distinguished baseline HCC samples, follow-ups with recurrence, follow-ups without recurrence, and controls. Baseline HMS>0.9 was associated with higher recurrence risk in Cox regression (HR 3.43 (95% CI 1.30-9.06), p=0.013). In all patients with follow-up samples, HMS decreased by 3-44% (median 17%) within the first 13 weeks after surgery. Subsequently, HMS trajectory of recurrent and non-recurrent patients diverged, with HMS rise relative to the first post-surgery timepoint associated with clinical recurrence. HMS functioned independently of other clinicopathologic variables.

**Conclusion:** Tumor-agnostic cfDNA methylomes accurately detect HCC and predict recurrence after liver resection or transplantation. This approach may have important implications for HCC diagnosis, treatment, and monitoring.

## INTRODUCTION

Hepatocellular carcinoma (HCC) is the most common primary liver malignancy and the third most common cause of cancer-related death worldwide. The overall 5-year survival rate for HCC remains less than 15-20%, as the majority of patients have advanced disease at diagnosis and do not qualify for potentially curative therapies [1]. Novel immunotherapy protocols have begun to show improved outcomes not only as palliative treatment for advanced HCC but also as adjuvant and neoadjuvant treatment in surgical patients [2]. Patients with HCC confined to the liver, preserved liver synthetic function and no portal hypertension are eligible for surgical resection, while those with decompensated liver disease or portal hypertension can undergo liver transplantation if deemed fit for major surgery. Despite curative-intent surgery, however, up to 60-70% of resection patients and 20-30% of transplant patients experience HCC recurrence within 5 years [3, 4].

Definitive diagnosis of de novo or recurrent HCC requires contrast-enhanced CT/MRI or biopsy, though the latter may carry significant risks in patients with decompensated liver disease [5]. While an elevated serum α-fetoprotein (AFP) level has a high specificity for HCC in adults, it lacks sensitivity and is not elevated in a significant proportion of patients with HCC [6]. HCC staging and treatment decisions are currently based predominantly on tumor burden measured by imaging. The development of novel minimally invasive approaches to the assessment of HCC tumor biology using molecular and genomic techniques could facilitate HCC diagnosis, better guide patient selection for various treatment modalities, and improve outcomes for HCC patients. In recent years, analysis of circulating tumor DNA (ctDNA, tumor-derived cell-free DNA (cfDNA)) in the peripheral blood plasma of patients with cancer has been studied for its ability to detect disease and monitor progression or treatment response. The success of this approach, however, largely relies upon knowledge of the mutational profile of the underlying primary tumor. In HCC, the practicality of ctDNA sequencing for early disease detection is limited by frequent tumor multifocality and wide mutational heterogeneity that may evade the scope of tumor-naïve targeted gene panels [7]. The feasibility of cfDNA sequencing in HCC has largely been demonstrated in patients with advanced disease where tumor tissue is available [8–11]. In contrast to variations at specific genetic loci, methylation of DNA at CpG sites is a key determinant of cellular identity that may evolve with the development of disease, allowing for the identification of tissue-specific cancer-associated methylation patterns [12]. Although the clinical utility of specific methylated DNA markers in plasma is being investigated in HCC [13, 14], the use of genome-scale methylation profiling as a diagnostic, prognostic and surveillance tool in HCC has yet to be described. Cell-free methylated DNA immunoprecipitation and high-throughput sequencing (cfMeDIP-Seq) is an approach that is not constrained by the requirement to know specific tumor mutations, instead examining cancer-associated DNA methylation signatures across the genome [15]. cfMeDIP-Seq has been successfully used to detect a variety of other cancers noninvasively, in a tumor agnostic fashion, and with minimal requirement for sample input [15–18].

We sought to develop a minimally invasive, tumor-agnostic cfDNA assay for HCC that could detect minimal disease and monitor disease progression. Patients with an established diagnosis of HCC undergoing surgery aimed at clearing all detectable disease, but with a significant risk of postoperative recurrence, provided an ideal study population. Here we report the first application of cfMeDIP-Seq to HCC for detection and monitoring of recurrence after curative-intent liver resection or transplantation.

## RESULTS

### Patient cohort, clinical characteristics, and blood collections

Of 92 patients recruited, 3 were excluded due to failed cfDNA isolation or QC analysis. Table 1 and Fig. 1 summarize the demographic and clinical data of the 89 HCC patients enrolled into the study. 32 (36.0%) underwent liver resection and 57 (64.0%) received a liver transplantation. For patients who had resection, surgery was the first treatment received in the majority of patients. In contrast, the majority of those undergoing liver transplantation received locoregional bridging therapies aimed at controlling HCC progression during their time on the waitlist. HCC recurred in 20 of 32 (63%) patients who underwent resection, which was significantly more common than 13 of 57 (23%) patients who underwent transplantation. Greater tumor burden and presence of microvascular invasion were also associated with recurrence (Table 1). Comparison of patients who underwent resection as compared with those who underwent liver transplantation revealed expected differences with regards to tumor stage, size, number and focality (Supplementary Table 1).

**Fig. 1.**
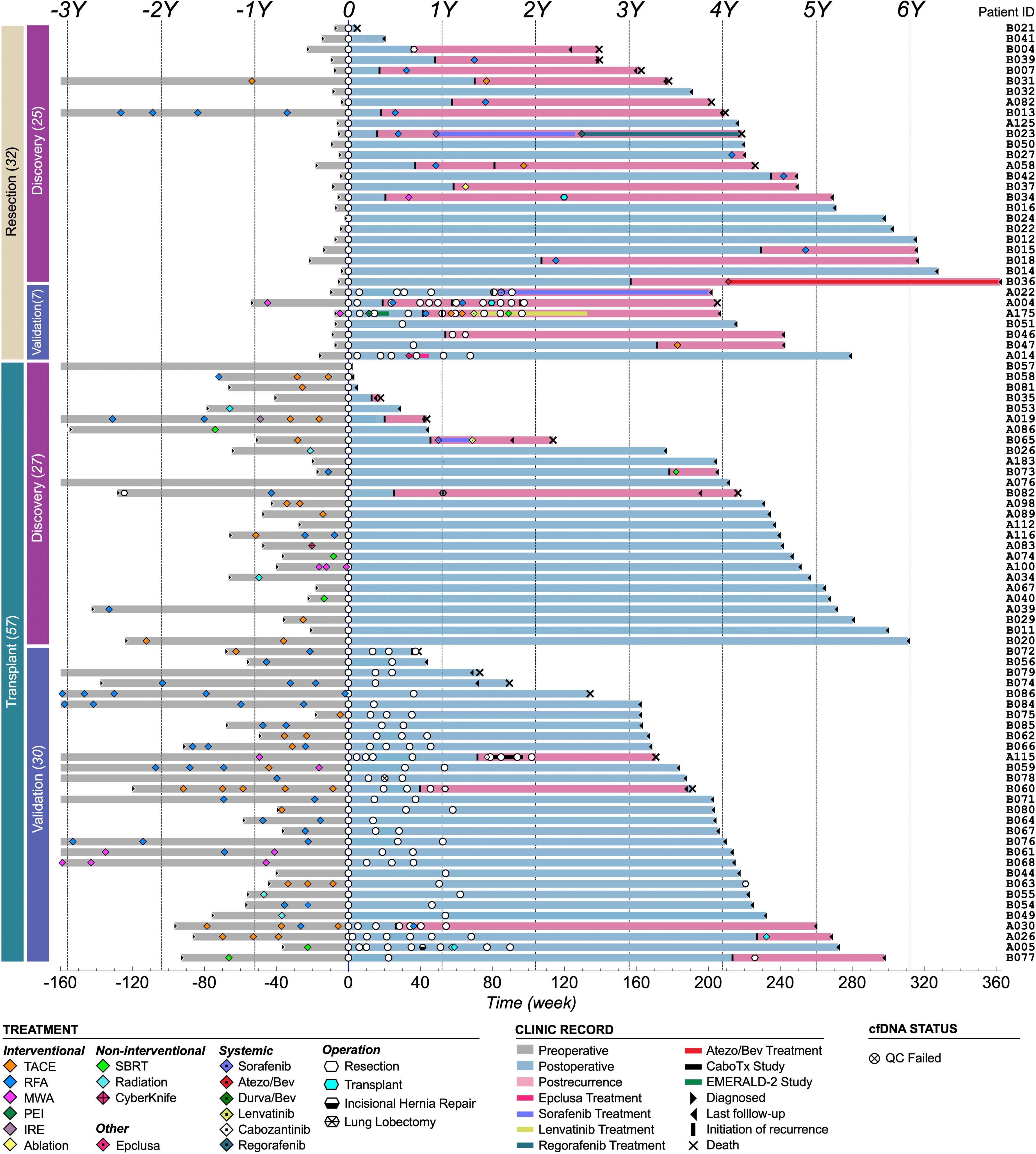
Clinical course of patients in study cohort. Swimmer plot illustrating the clinical course of patients in the study grouped by type of surgery (resection or transplant) and machine learning cohort (discovery or validation). The day of surgery is designated as time 0. The X-axis represents the timeline in week. The preoperative timeline (up to three years before surgery) is displayed as a gray bar, while the postoperative timeline is shown as a light blue bar. For patients developed recurrence, the post-recurrence timeline is depicted in pink. Time points in years are marked along the top with corresponding vertical lines. Plasma samples collected for cfMeDIP-Seq are represented by white circles. Patient IDs are labeled on the far right, legends for other symbols are included below. Source data are provided as supplementary file.

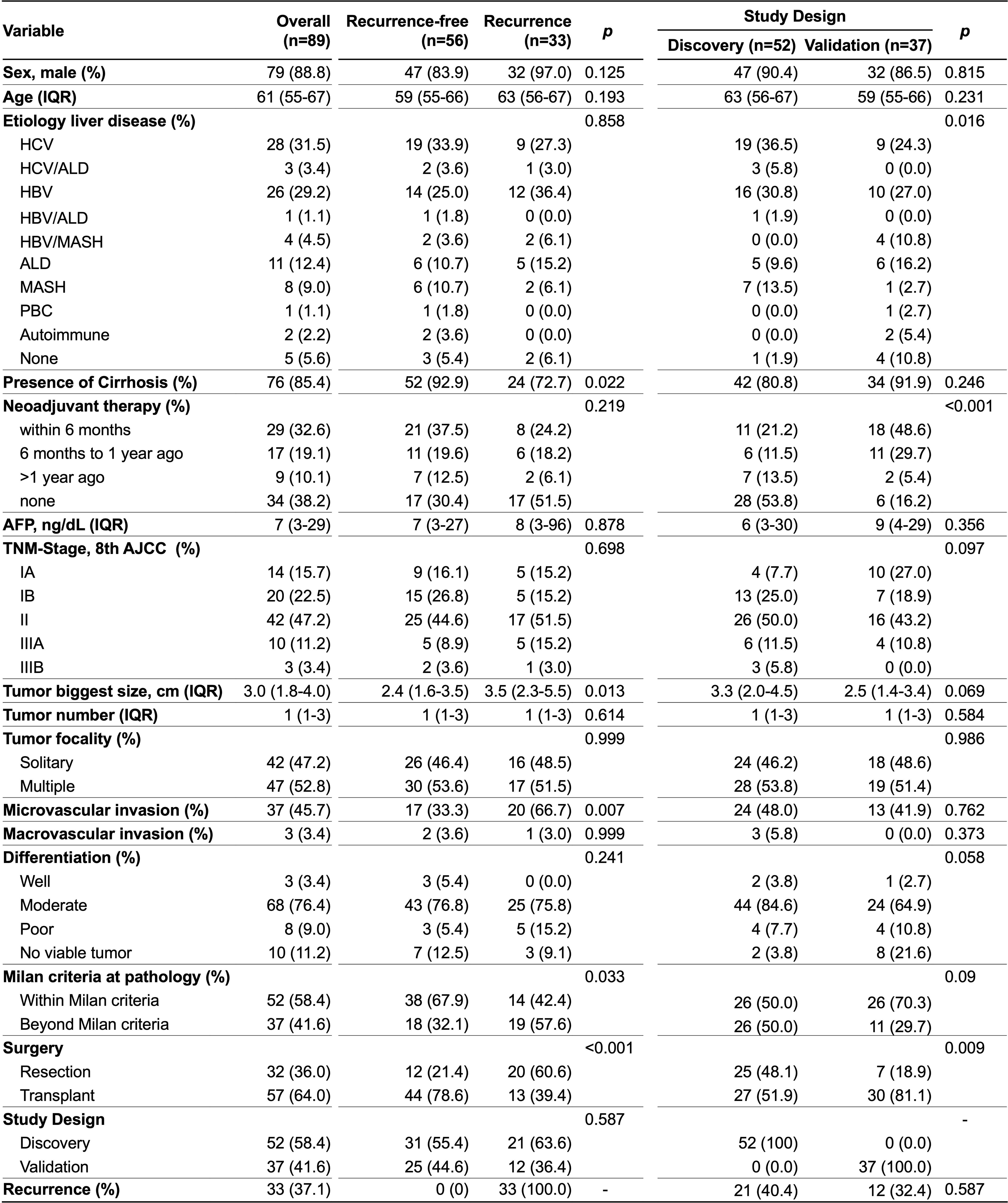

A total of 124 baseline plasma samples were obtained on the day of surgery from all 89 HCC patients (b-HCC) and from 35 healthy controls (CTL). From this population, we defined a discovery cohort including 35 CTL and 52 b-HCC samples from whom we only had baseline but no follow-up blood collections. The remaining b-HCC samples from 37 patients and 112 corresponding follow-up blood samples (f-HCC) collected at subsequent clinic visits were included in a validation cohort (Fig. 1, Fig. S1). Among the total of 112 f-HCC samples, 25 were collected at various time points after the diagnosis of recurrence from 8 patients. These include 10 samples from 5 transplant cases and 15 samples from 4 resection cases. The remaining 87 samples consisted of 21 pre-recurrence postoperative samples from 7 patients who ultimately developed HCC recurrence, as well as 66 samples from 30 patients who remained recurrence-free throughout the follow-up period.

As shown in Table 1, clinical variables were identical between discovery and validation cohorts, except for an over-representation of patients receiving bridging therapy which was more common in the validation cohort due to the higher proportion of transplant patients in this group who were more likely to receive pre-transplant locoregional therapy. Because we used the availability of postoperative follow-up blood samples as a criterion to determine the makeup of the discovery and validation cohorts, the proportion of transplant patients in the validation cohort was greater than in the discovery cohort due to the greater frequency with which transplant patients presented to clinic for routine follow-up and bloodwork.

Blood was also collected at a single time point from a total of 35 healthy controls. These individuals included 23 living liver donors just prior to their donor hepatectomy (41.7% male, mean age: 35.5). Living liver donors were confirmed to be free of cancer or liver disease through an extensive standard panel of laboratory tests and diagnostic imaging required for living donation. We combined these data with cfMeDIP-seq sequencing data from another cohort of 12 healthy blood controls from other studies. [18].

### Development of HCC classifier and HCC methylation score (HMS)

To establish a set of top differentially methylated regions (DMRs) for scoring the presence of HCC-derived cfDNA, we compared 52 b-HCC blood samples with 35 healthy controls (CTL) in the discovery cohort using a 10-fold cross-validation machine learning strategy. For each of 100 training-testing iterations, the top 300 DMRs individually yielded a mean AUROC of 0.999 (95% CI 0.998 – 1; Fig. 2A). By intersecting the top 300 DMRs from each of the 100 iterations, we obtained a final set of 4994 unique sites. These were then utilized for principal component analysis (PCA) to compare the baseline discovery cohort with the healthy controls. This consensus set of DMRs demonstrated clear separation of the healthy controls from both the discovery b-HCC (Fig. 2B) and the validation cohort (Fig. 2C). When extended to the full cohort including follow-up samples, all b-HCC (yellow) and f-HCC samples diagnosed with recurrence (f-HCC-Rec, red) clustered together distinctly from healthy controls (Fig. 2D). In contrast, f-HCC samples diagnosed without recurrence (remission, f-HCC-Rem, blue) did not cluster tightly and were interspersed with healthy controls and b-HCC samples.

**Fig. 2.**
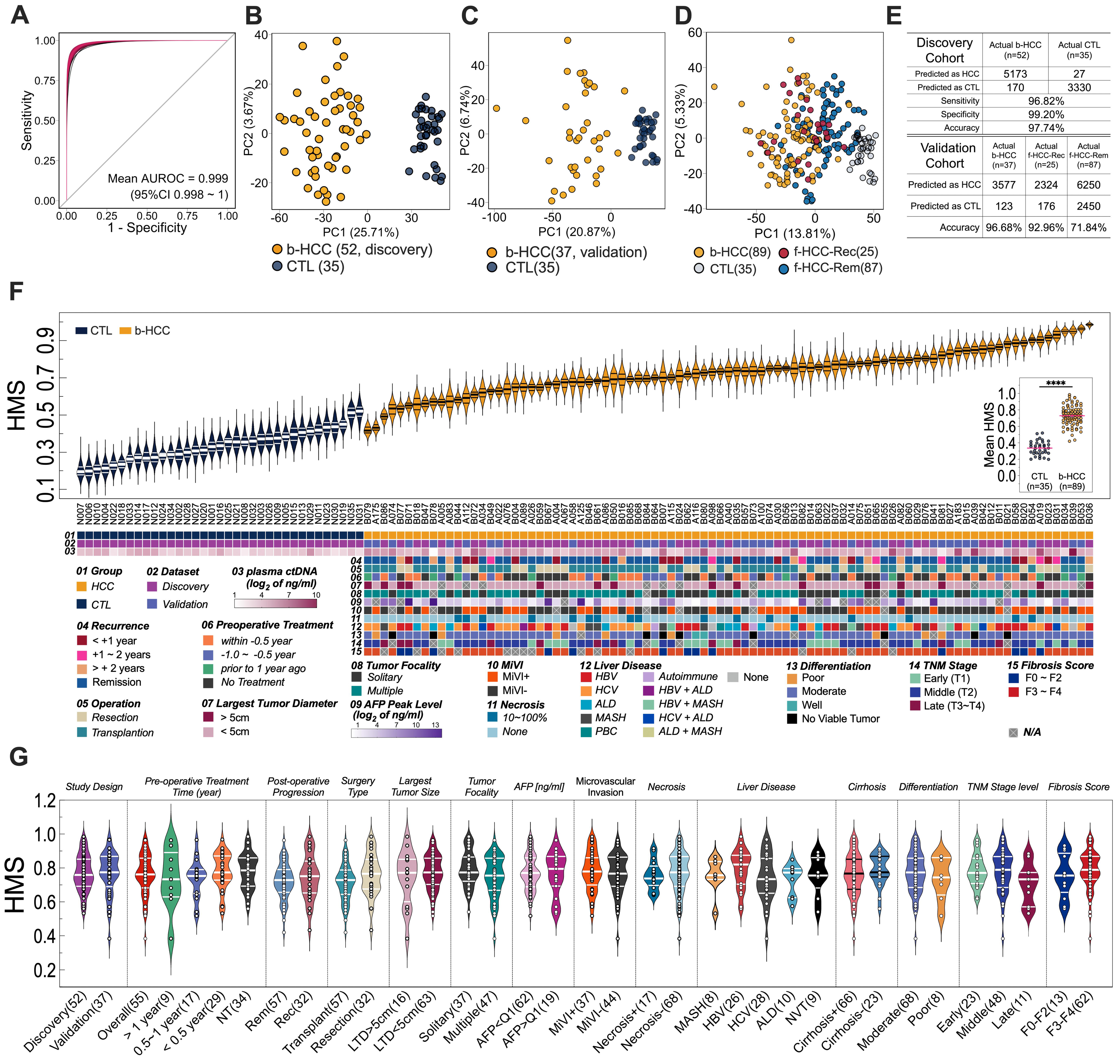
Identification of HCC using ctDNA methylomes. (A) Overlaid ROC curves for 100 iterations of b-HCC (n=52) vs. CTL (n=35) classifiers. (B-D) Principal component analysis (PCA) plots for cohort of (B) baseline discovery, (C) baseline validation and (D) all samples generated using the set of 4994 HCC-specific DMRs identified during training-testing iteration repeats. (E) Confusion matrix showing the sensitivity, specificity, and accuracy of modeling performance of discovery cohort (top) and prediction accuracy (predicted as HCC) of validation cohort samples. (F) Violin plots show raw HCC methylation score (HMS, top) across all b-HCC and healthy controls samples, inset scatter plot shows the mean of b-HCC and CTL mean HMS, heatmap below illustrates clinical data corresponding to each baseline sample. (G) Violin plots showing the HMS for all b-HCC samples grouped by different clinical parameters. (F,G) Violin plots indicate median ± IQR, scatter plots indicate mean ± SE. Source data (for panel F and G) provided as supplementary file. SE, standard error; LTD, largest tumor diameter; MiVI, microvascular invasion.

The performance of the discovery cohort-based HCC classification model is summarized in the confusion matrix (Fig. 2E, top), with an extremely high sensitivity of 96.82%, specificity of 99.20%, and accuracy of 97.74%. In the validation cohort (Fig. 2E, bottom), the classifier identified 37 b-HCC samples with an accuracy of 96.68%. The accuracy of the classifier for f-HCC-Rec samples was 92.96%. However, the classifier correctly identified f-HCC-Rem samples only 71.84% of the time, suggesting greater variability in the detection of HCC remission and a more challenging task for the classifier in distinguishing these cases.

To establish a probabilistic HCC Methylation Score (HMS, range 0-1), we applied the exact 100 list of top 300 DMRs to the validation cohort which included 37 (longitudinal) b-HCC and 112 corresponding f-HCC samples. This group design maintains the absolutely consistent parameters when generating HMS for b-HCC and f-HCC collected from the same patient and guarantees the reliability of subsequent results obtained through pairwise strategies for post-surgery HMS trajectory analysis.

As shown in Fig. 2F, according to the outcome of 100 training-testing repeats, 87 of 89 (97.75%) baseline samples from both discovery and validation cohorts were assigned a higher HMS than all healthy control samples. The overall mean HMS for the b-HCC samples was significantly higher than healthy controls (0.767 vs. 0.336) (inset scatter plot). Notably, we evaluated potential correlations between HMS and an extensive collection of other clinically relevant variables (depicted for each sample in Fig. 2F heatmap) but found no significant results (Fig. 2G).

### HCC sub-typing by cfMeDIP-Seq

We then employed a “one vs each” tumor subtyping machine learning strategy [16] to determine whether DMRs identified through cfMeDIP-Seq could differentiate HCC subtypes at baseline arising on different chronic liver disease backgrounds. As shown in Fig. S3A, this approach allowed discrimination of HBV-related HCC from healthy controls and HCC arising on other backgrounds (mean AUROC = 0.916, 95% CI = 0.904-0.938). Similarly, PCA plots in Fig. S3B reveal clustering of samples from HCC patients according to underlying liver disease etiology and distinct from healthy controls. Clusters are most pronounced for HCC arising on a background of HBV, HCV and ALD with no clear distinction between cirrhotic and non-cirrhotic cases within each subgroup. To exclude the possibility that the presence of cirrhosis was confounding the DMR signal being attributed to the presence of HCC, we compared the HMS values assigned to HCC samples arising on a background of cirrhotic vs. non-cirrhotic HBV-related liver disease. As shown in Fig. S3C, no difference was detected, strongly confirming that HCC-specific DMR signatures identified in our analyses are related specifically to cancer rather than other manifestations of chronic disease in the liver.

### Baseline HMS and postoperative HMS trajectory predict HCC recurrence

We next assessed the utility of HMS at baseline and during postoperative follow-up to predict HCC recurrence. Despite potential differences in the pathophysiology of HCC recurrence following resection and transplant (Fig. 3C), HMS did not distinguish between resection and transplant samples at baseline(Fig. S4, A, E), and ΔHMS also demonstrated minimal to no distinction during follow-up.(Fig. S4, B-D, F, G). To strengthen our analysis in light of the small number of patients involved, we analyzed HMS by recurrence or remission status in aggregate without further stratifying them by resection or transplant status.

**Fig. 3.**
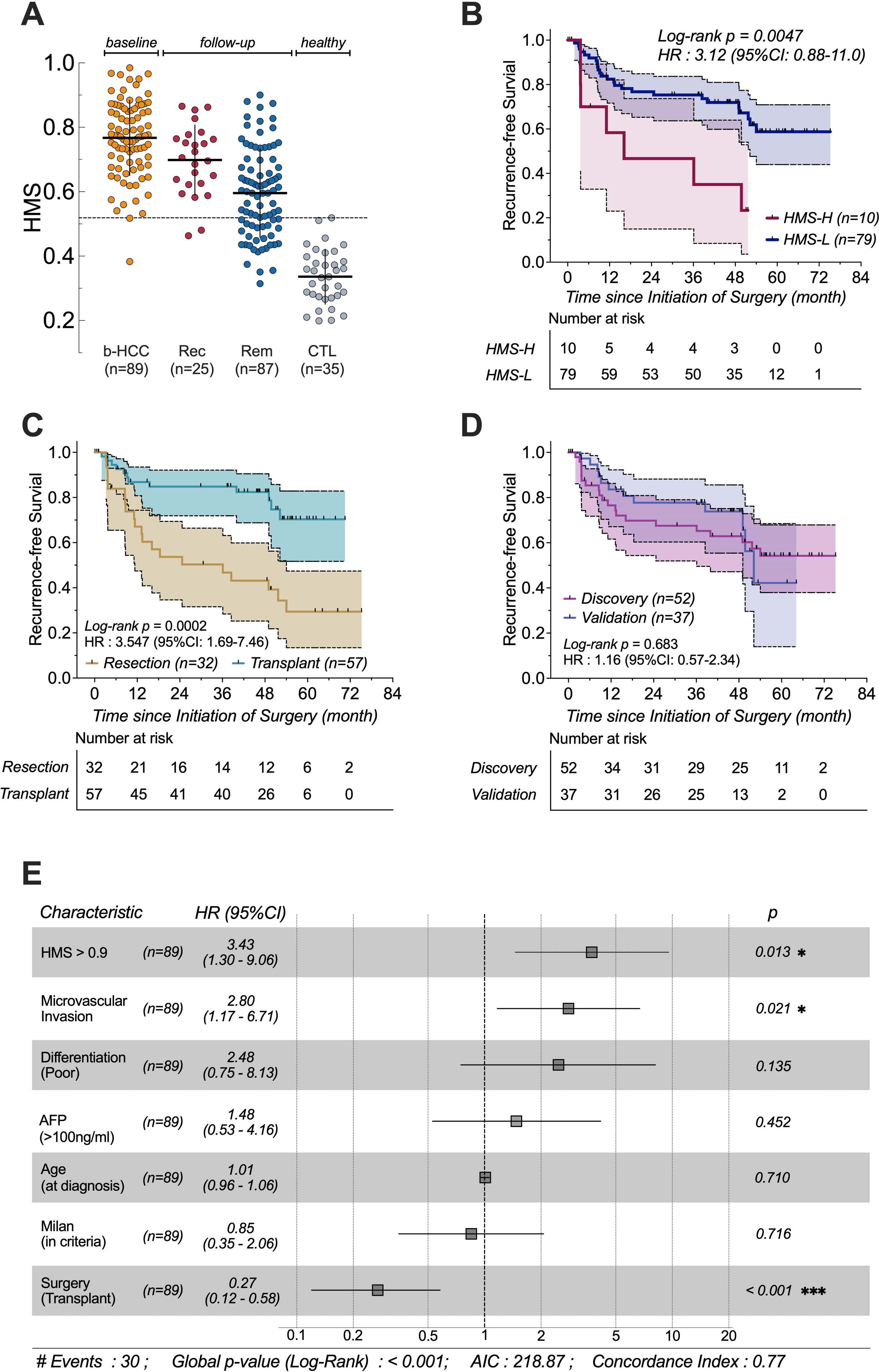
Recurrence-free Survival (RFS) analysis. (A) Scatter plots (mean ± SD) depicting the HMS value across all tested individual patients in four groups, including 89 baseline HCC (b-HCC), 112 follow-up HCC diagnosed with (Rec, n=25) or without recurrence (remission, Rem, n=87) and 35 healthy control (CTL) samples. Horizontal dashed line indicates the highest HMS value (0.519) of healthy controls. Significant difference (p < 0.01, two-tailed Mann– Whitney U test) was observed in each pairwise comparison between two groups. (B-D) Kaplan-Meier curves of RFS between patients with HMS > 0.9 and HMS < 0.9 (B), received liver resection or transplantation (C) and designated discovery or validation cohort (D). (E) Multivariate Cox proportional-hazard regression analysis of associations between clinicopathologic characteristics and HCC patient survival outcomes. * p□<□0.05, *** p□<□0.001. HR, hazard ratio; SD, standard deviation. Source data (for panel A and B) provided as supplementary file.

**Fig. 4.**
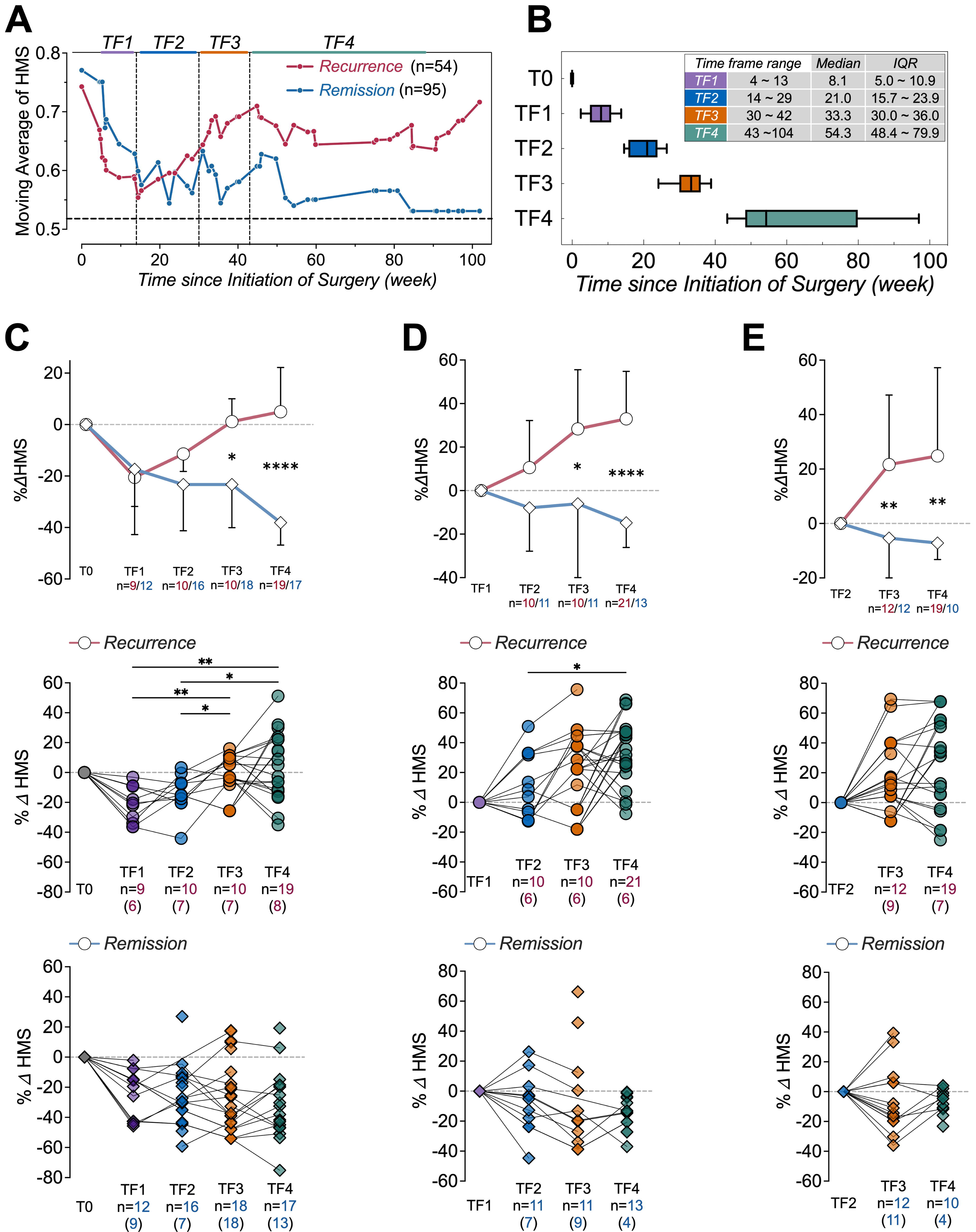
Postoperative HMS trajectory and HCC outcome. (A) Moving average of HMS values in recurrence and remission patients beginning at time of surgery and various time points during postoperative follow-up. Horizontal dashed line indicates the top HMS value (0.519) of healthy control. (B) Box and whiskers plot illustrates the distribution of 4 timeframes (TF) based on the inflection and divergence points in moving average curves. Box represents median ± IQR, whiskers extend to the 5-95 percentiles, capturing 90% of the data distribution. Detailed information about the time span and statistical results is shown in the top right corner. (C-E) Percent change in mean HMS values for recurrence and remission patients during postoperative timeframes. Graphs at top illustrate percent change (%ΔHMS) in overall mean HMS values for all patients in each group normalized to T0 (C), TF1 (D), or TF2 (E) with error bars indicating median ± IQR. Rows of graphs below (Recurrence at middle, Remission at bottom) show the %ΔHMS at single data point resolution to display the dispersion. n indicates the number of cfMeDIP tests with the number of patients from whom these samples were sourced indicated in parentheses below. * p < 0.05, ** p < 0.01, **** p < 0.0001. IQR, interquartile range. Source data provided as supplementary file.

As shown in Fig. 3A, the comparison of HMS values among baseline HCC (b-HCC), follow-up (f-HCC) samples diagnosed with recurrence (Rec) and without recurrence (Rem), and healthy controls (CTL) showed significant differences. Specifically, the highest HMS value among 35 healthy control samples is marked by a horizontal dashed line at 0.519. Notably, only 2 of 89 b-HCC and 2 of 25 recurrence f-HCC samples have HMS values lower than this threshold. In contrast, 28 of 87 (∼32%) remission f-HCC samples fall below this threshold, aligning their values with those seen in healthy controls.

We then evaluated the relationship between HMS values at baseline and recurrence-free survival (RFS). As shown in Fig. 3B, RFS was significantly reduced in patients once HMS exceeded 0.9 at baseline. Furthermore, as demonstrated in Fig.3C, Cox multivariable regression analysis revealed that HMS > 0.9 was associated with an increased risk of recurrence with a hazard ratio of 3.43 (95%CI 1.30-9.06, p=0.013). However, baseline samples exceeding an HMS threshold of 0.9 represented only a small fraction of the cohort (10 of 89 patients), making this threshold of limited clinical relevance. RFS analysis results also showed no significant difference between the Discovery and Validation cohorts (Fig. 3D), suggesting the reliability of our machine learning group design.

We next examined whether trends in HMS trajectory after surgery were associated with HCC recurrence. Due to variability in post-operative sampling times between patients, we first determined the temporal trend in mean postoperative HMS for patients with and without recurrence using a simple moving average method (Fig. 4A). By identifying critical divergence points in these trends, we divided the postoperative sampling period into four timeframes (Fig. 4B): TF1 (5.0-10.9 weeks), in all patients, HMS decreased by 3% to 44% (median 17%) within the first 13 weeks after surgery. TF2 (15.7-23.9 weeks), HMS began to increase in recurrent patients. TF3 (30.0-36.0 weeks), HMS in recurrent patients continued to rise towards baseline levels, while significant fluctuations were observed in non-recurrent patients. TF4 (48.4-79.9 weeks), recurrent patients maintained high HMS levels, whereas HMS consistently declined in those without recurrence. The median times of these timeframes are 8.1 (TF1), 21.0 (TF2), 33.3 (TF3), and 54.3 (TF4) weeks after the surgery. Next, we determined the change in HMS at each sampling point within consecutive timeframes for each patient (Fig. 4, C-E). As shown in Fig. 4C, the mean HMS decreased in all patients between surgery and first follow-up. Subsequent to this, the mean HMS increased in patients with recurrence while it stabilized or further decreased in those who did not recur. Interestingly, in those who developed recurrence, the absolute mean HMS did not return to baseline levels until the third sampling time frame (TF3). The clearly divergent patterns in mean HMS change between recurrent and non-recurrent groups were more clearly evident when plotted relative to the mean HMS at the first (TF1) and secondary (TF2) postoperative follow-up timeframes, as shown in Fig.4D and 4E.

Having observed that aggregate HMS trajectories distinguish patients with recurrence from those without, particularly after first follow-up postoperative follow-up timeframe (TF1), we examined HMS during follow-up at single-patient resolution to determine how closely HMS reflects HCC recurrence or progression. Fig. 5 illustrates percent change in HMS (%ΔHMS) during the postoperative course of nine representative patients relative to TF1, three with HCC recurrence in four patients who underwent liver resection (Fig. 5A). And three with HCC recurrence in six patients who underwent liver transplant (Fig. 5B). In general terms, these plots demonstrate that in patients who go on to develop recurrence, the HMS remains at or above the baseline reached at the first postoperative follow-up, and all patients with recurrence eventually exhibit a significant HMS increase (ranging from 22% (A022) to 49% (A030)). In these patients, %ΔHMS temporally coincides with clinical evidence of HCC recurrence and decreases following tumor-directed therapies. In contrast, in patients who demonstrate sustained remission of disease after surgery, %ΔHMS continues to drop below the baseline value achieved at the first postoperative follow-up sample, experiencing a substantial decrease ranging from 8.6% (A005) to 27.4%(A014). Notably, in nearly all patients, the key divergence points in HMS trends (Fig. 4D) indicating substantial increases (vertical crimson dash line) or decreases (vertical blue dash line) manifested earlier (or at least no later) than the times confirmed by clinical imaging diagnoses (vertical orange/black lines).

**Fig. 5.**
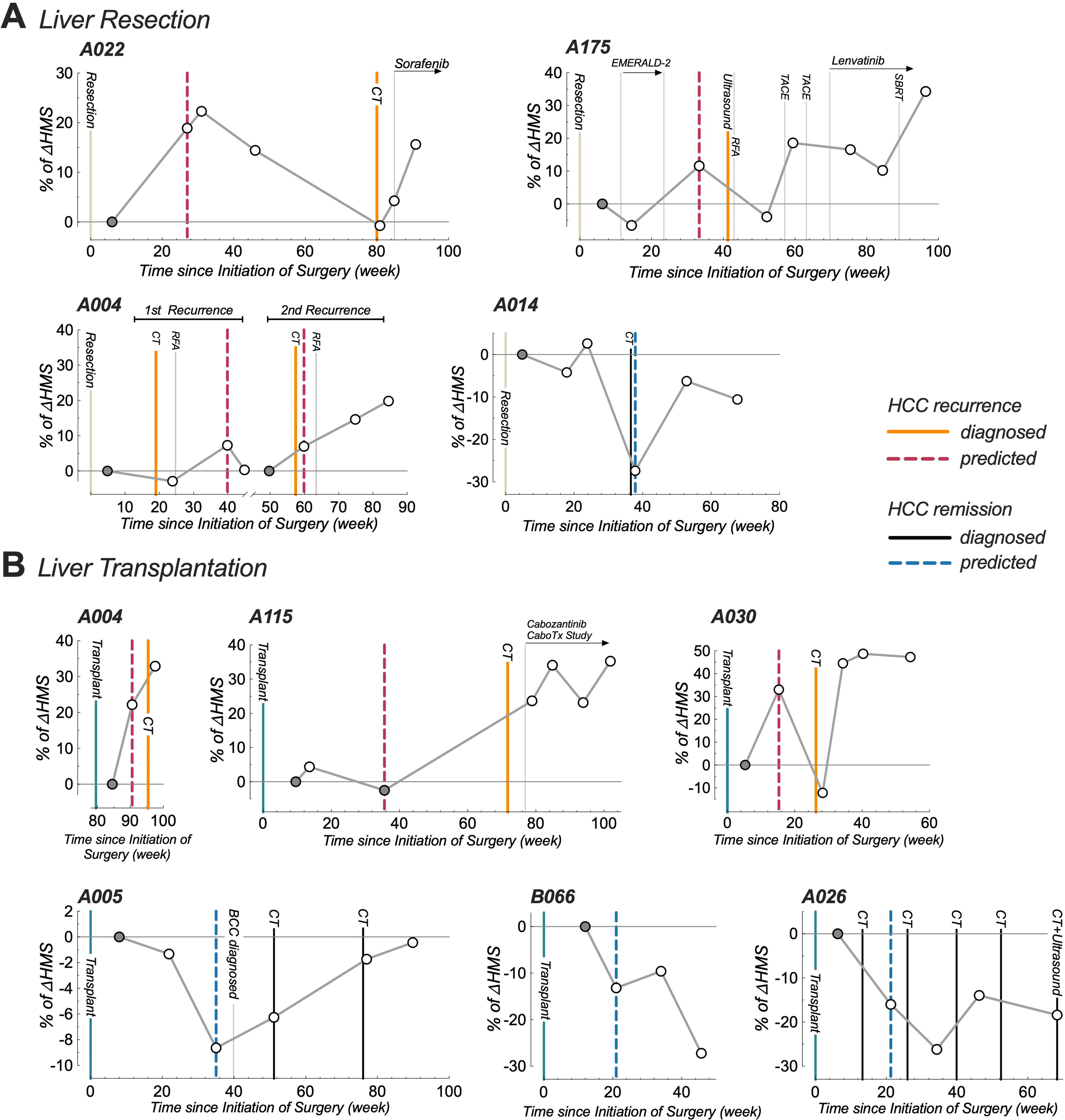
Postoperative HMS plots reflect disease course in individual patients. Longitudinal plots for 9 patients depicting the percent change in HMS (%ΔHMS) at various ctDNA collection time points (white circles) during postoperative follow-up normalized to TF1 (grey circle). Patients received liver resection (A) and liver transplantation (B) were shown separately. Grey horizontal line indicates the normalized baseline TF1 HMS, bold vertical lines indicate clinically confirmed postoperative recurrence (orange) or remission (black) events by imaging (CT or ultrasound). Bold vertical dash lines indicate the earliest time points for ΔHMS-predicted postoperative recurrence (crimson) or remission (blue) events, based on matching the trends to the earliest key divergence points identified in Fig. 4D. Other postoperative clinical treatments are marked by thin vertical grey lines with intervention labels. Patient ID is indicated at the top left of each panel. Source data provided as supplementary file. BCC, basal cell carcinoma.

For example, ΔHMS-predicted patient A175 recurrence was identified at 33.3 weeks, compared to 41.3 weeks as confirmed by CT; patient A022 at 27.0 weeks vs. 80.0 weeks; patient A030 at 15.3 weeks vs. 26.3 weeks; patient A115 at 35.6 weeks vs. 71.7 weeks; and patient A005 at 35.0 weeks vs. 51.1 weeks.

## DISCUSSION

In this study, we have demonstrated that cfMeDIP-seq, a tumour-agnostic, genome-wide methylation-based approach that enriches tumor-specific methylated cfDNA fragments, is capable of detecting HCC and monitoring recurrence after curative-intent surgical therapy. While others have reported liquid biopsy-based cfDNA technologies [19], many of these are challenged by the low abundance and high degradation of tumor-specific cfDNA, leading to false positives and poor test reliability [15, 20]. Furthermore, compared to existing commercial ctDNA-based diagnostic technologies, cfMeDIP-seq demonstrates superior features by requiring lower input cfDNA, performing with higher sensitivity and specificity [58–60], and offering broader applicability due to its tumor-agnostic nature [19, 50, 61–63]. According to recently published ctDNA-based HCC early detection studies [51, 52], our testing requires lower input cfDNA and performs with higher sensitivity compared to other methodologies.

While most prior studies of ctDNA monitoring in HCC included patients with advanced disease eligible for palliative therapies with limited prognosis, we focused on earlier-stage patients eligible for curative surgery or liver transplantation. Unlike many studies that target specific liver disease backgrounds, our diverse North American cohort represents the full spectrum of liver disease etiologies leading to HCC. We confirmed that cfMeDIP-seq can detect HCC with high sensitivity and specificity, unaffected by clinicopathologic variables, even in patients receiving bridging therapy with minimized tumor burden. Importantly, the developed HMS functioned independently of common HCC predictive variables including AFP levels, tumor number, size, and microvascular invasion (Fig 2F, 2G). These findings suggest cfMeDIP-seq is a valuable adjunct for HCC clinical management.

Beyond detection, our data demonstrate that a quantitative score system (HMS) generated by machine learning can predict post-surgery outcomes. While baseline HMS thresholds were predictive of recurrence (Fig 3B), the %ΔHMS trajectory appears to be more effective. Key HMS trend divergence points (Fig 4D), indicating significant increases or decreases, often appear earlier than conventional imaging diagnoses (Fig 5).

Interestingly, %ΔHMS compared to the first postoperative samples proved most useful. This suggests that an HCC epigenomic signature persists in plasma for weeks after curative therapy and can be detected by high-resolution sequencing technologies. Factors determining whether persistent microscopic disease evolves into clinically evident recurrences remain to be defined by future cfDNA studies.

Highly sensitive tumor-agnostic molecular detection of HCC using cfMeDIP-seq has a number of potential advantages over current clinical standards that rely on cross-sectional imaging and AFP. Earlier detection of microscopic recurrent disease using cfDNA would offer an opportunity to improve outcomes by initiating adjuvant therapies [22], modifying immunosuppression in transplant recipients, and tailoring the use of surveillance imaging. Our data also suggest that cfMeDIP-Seq should be explored as a novel method to stratify post-transplant HCC recurrence risk of liver transplant candidates; if effective, this would represent a much-needed biomarker that could be incorporated into transplant listing and organ allocation decisions to maximize the utility of scarce donor organs.

One limitation of this study is the small size of the cohort. Despite the substantial HCC cohort of 112 postoperative follow-up samples collected from 37 patients, only 15 underwent cfMeDIP-seq testing at least three times. These patients, ideal for longitudinal trajectory analysis, include individuals both with (n=7) and without (n=8) diagnosed postoperative HCC recurrence. Despite these relatively small groups, the HMS scores were highly divergent early in the follow-up timepoints demonstrating strong differences in cfDNA trajectory between post-surgery recurrence and non-recurrent HCC patients. Encouragingly, in the five distant relapse cases, HMS increases were observed at least 10 weeks before CT confirmation (Fig 5A, B). Despite potential pathobiology differences in HCC recurrence after resection versus transplantation, our small sample size limited HMS’s ability to distinguish these groups. While a larger prospective study cohort is essential to validate our findings, it is noteworthy that several reports describing the use of cfMeDIP-seq for detection of other cancers also had limited cohort sizes [15–18, 53]. Furthermore, recent high-impact publications highlighting the effectiveness of other cfDNA-based cancer detection techniques have demonstrated impressive outcomes based on serial samples from relatively small numbers of patients [54–57].

In conclusion, cfMeDIP-seq can accurately detect HCC and predict recurrence after liver resection or transplantation. This novel tumor-naïve cfDNA methylomes profiling approach may have important implications for HCC diagnosis, treatment, and monitoring.

## Supporting information

Supplementary Files Description

Supplementary_file_1_Clinic_records

Supplementary_file_2_HMS_anaylsis_raw_data

Supplementary_file_3_QC_raw_data

Supplementary_file_4_HMS_trajectory_data

Supplementary_file_5_HMS_extended_analysis_data

---

**Supplementary Fig. S1.**
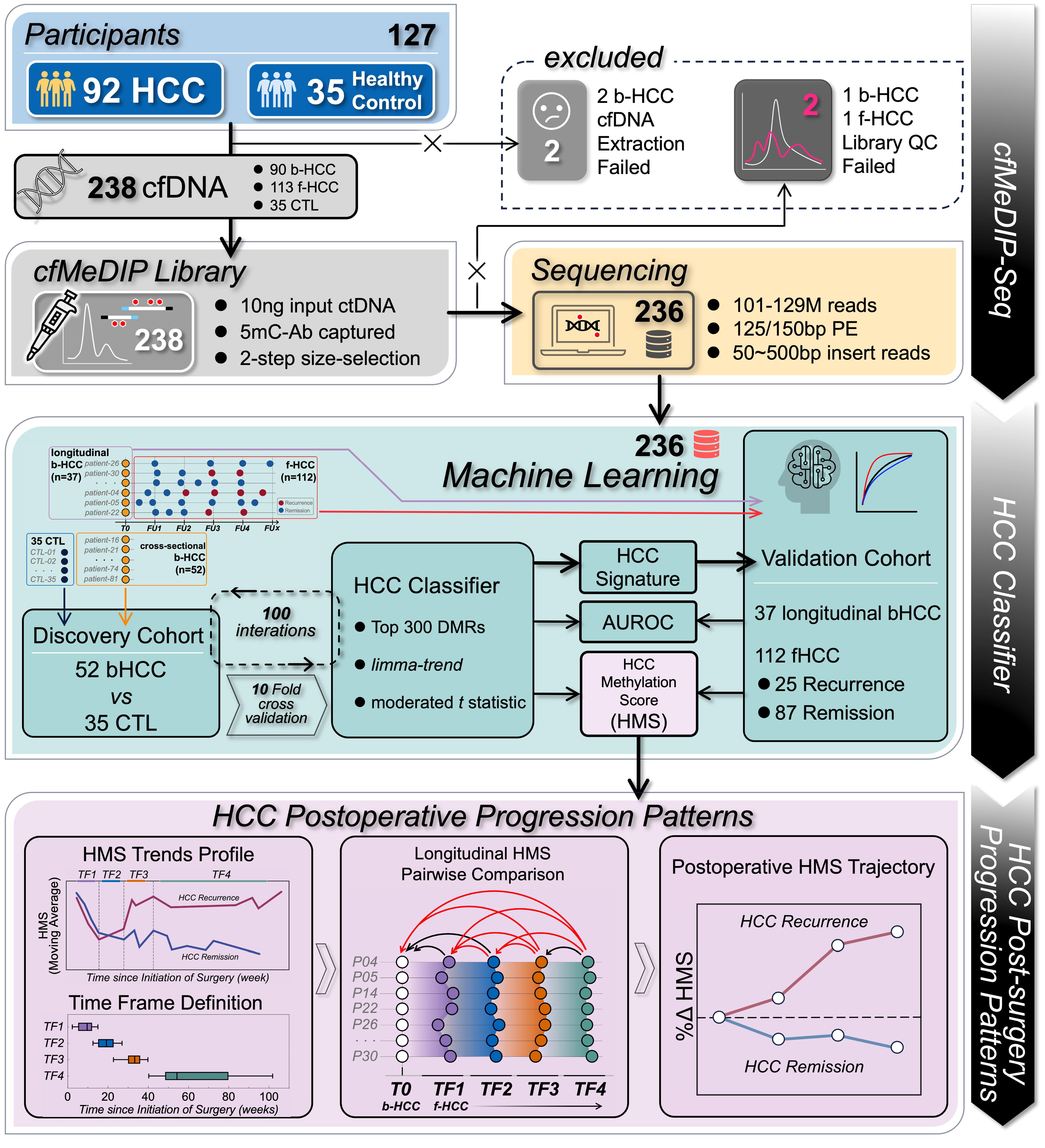
Schematic Flowchart of Study Design. Total of 240 ctDNA samples was successfully isolated from 238 plasma samples collected from 127 patients. cfMeDIP libraries were generated successfully in 236 of these samples and were sequenced (89 baseline HCC, 112 follow-up HCC, and 35 healthy controls). For development of the HCC classifier and HCC methylation score (HMS) using machine learning, the discovery cohort consisted of 35 healthy controls and 52 baseline HCC samples that did not have matched follow-up samples. The model was then tested on a validation cohort consisting of 37 baseline HCC samples and 112 matched follow-up samples. HMS values at baseline and during postoperative follow-up were then analyzed to determine their relationship with HCC recurrence and other clinicopathologic variables.

**Supplementary Fig. S2.**
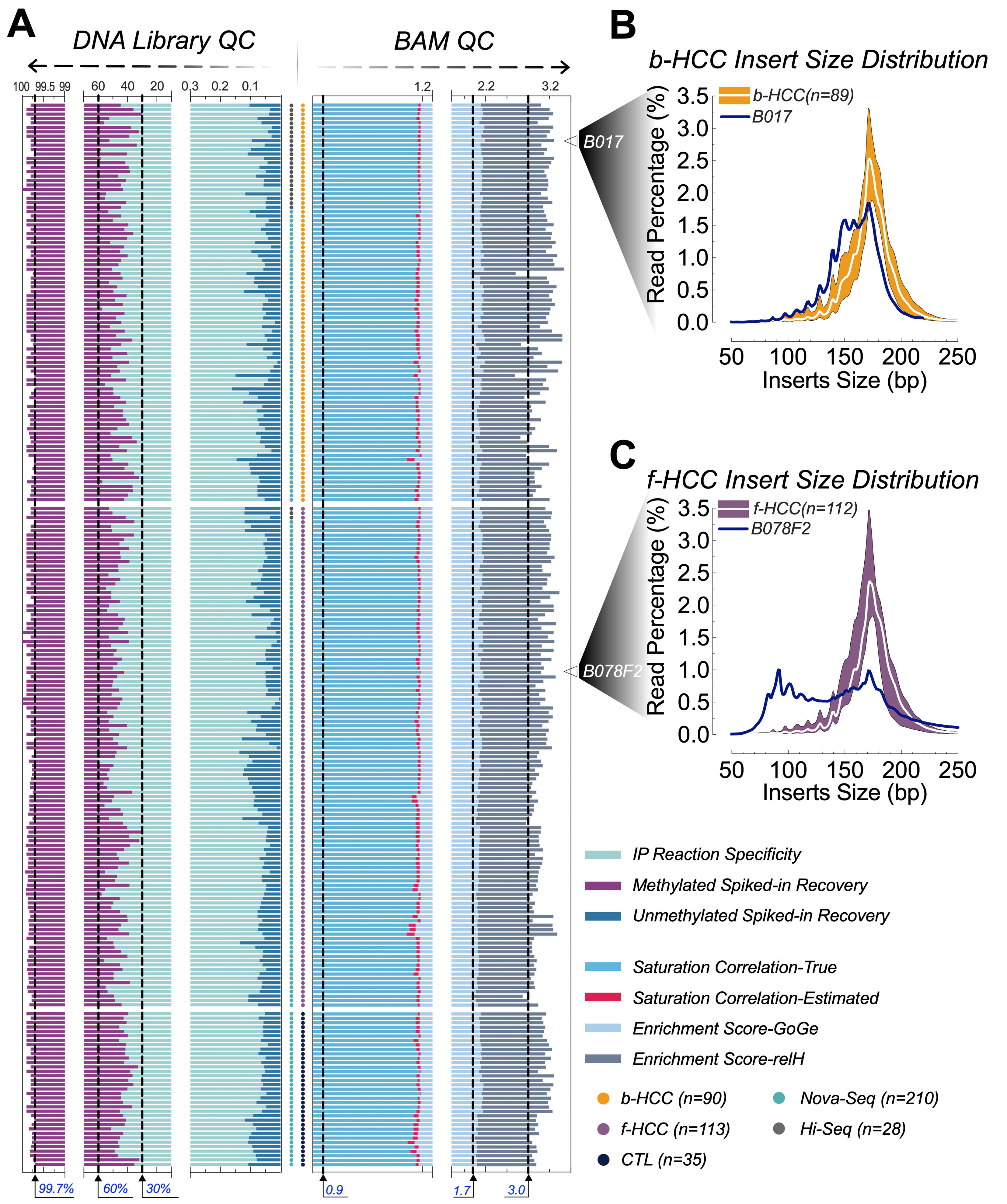
Quality control (QC) of cfMeDIP sequencing. (A) QC of cfMeDIP DNA libraries (left) and BAM files (right). Recovery and specificity of the 5mC antibody immunoprecipitation (IP) are calculated using the percentage recovery of methylated and unmethylated spiked-in A. thaliana DNA. cfMeDIP libraries that failed to meet all three key assessment indicators including 0.05-0.3% of unmethylated spiked-in DNA recovery, 30-60% of unmethylated spiked-in DNA recovery and 99.7-99.9% of IP reaction specificity [28] were reprepared. BAM files with reasonable CpG motif enrichment score (relH > 3.0 and GoGe > 1.7) and saturation (correlation or estimated correlation > 0.9) [28] were proceeded to downstream analysis. Sample groups and sequencing platforms are also indicated using circle dots. (B-C) ISD (Insert size distribution) analysis on FASTQ files generated from b-HCC (B) and f-HCC (C) cfMeDIP libraries within 50-bp to 250-bp size window. For b-HCC samples, the inner white line indicates the median of 89 FASTQ files, data range is highlighted in yellow. For f-HCC samples, the inner white line indicates the median of 112 FASTQ files, data range is high-lighted in purple. The abnormal ISD curves of two samples (1 b-HCC B017 and 1 f-HCC B078 secondary follow-up (B078F2)) which passed both QCs on cfMeDIP DNA libraries and BAM files is shown in dark blue. Source data provided as supplementary file.

**Supplementary Fig. S3.**
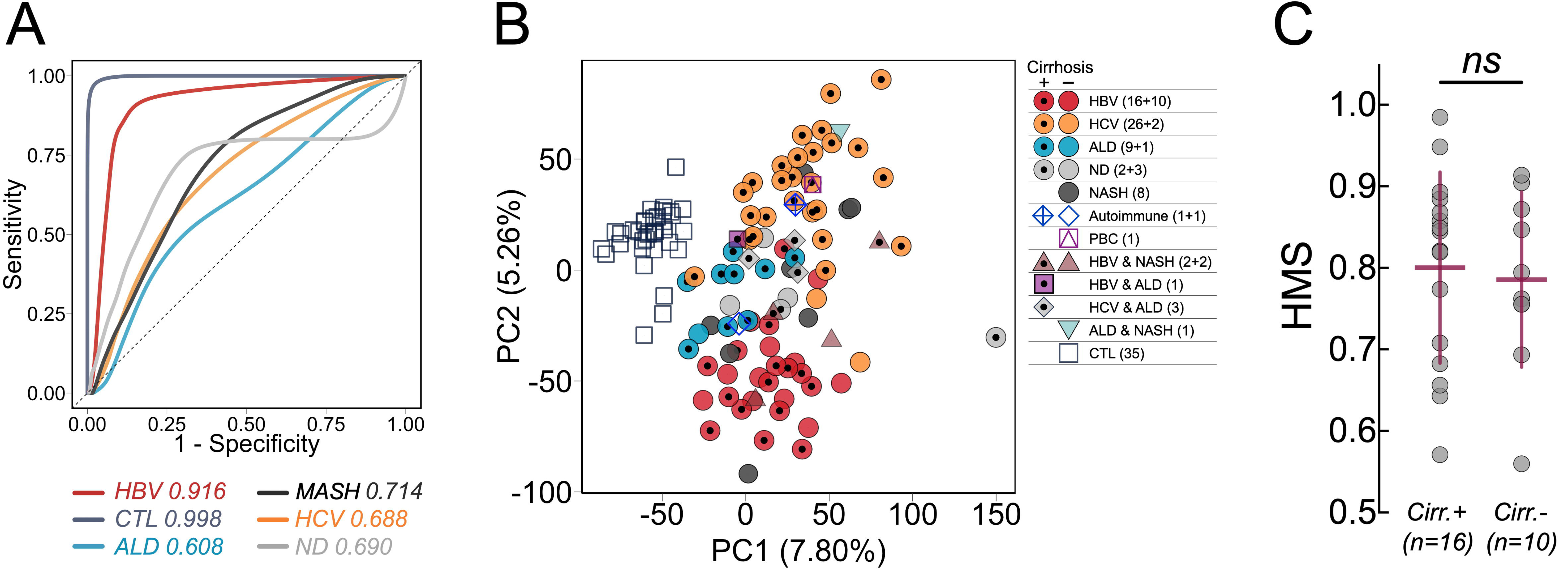
Identification of HCC subtypes by cfMeDIP-seq. (A) ROC curves constructed using averaged class probabilities for independent HCC sub-groups classified by different liver disease types including HBV (n=26), HCV (n=28), ALD (n=10), MASH (n=8), ND (n=5) and CTL (n=35) generated from 100 training-testing sets for one-class versus other-classes comparison trained using the full b-HCC cohort. (B) PCA plot generated using combined DMRs matrix for each vs. other classes model (2326 HBV-, 7976 HCV-, 3797 ALD-, 4104 MASH-, 3690 ND- and 4267 CTL-specific) (C) Scatter plot showing comparison between mean HMS for cirrhotic (Cirr.+, n=16) and non-cirrhotic (Cirr.-, n=10) samples within the HBV sub-group. Error bars represent mean ± SD. ns indicates no significance.

**Supplementary Fig. S4.**
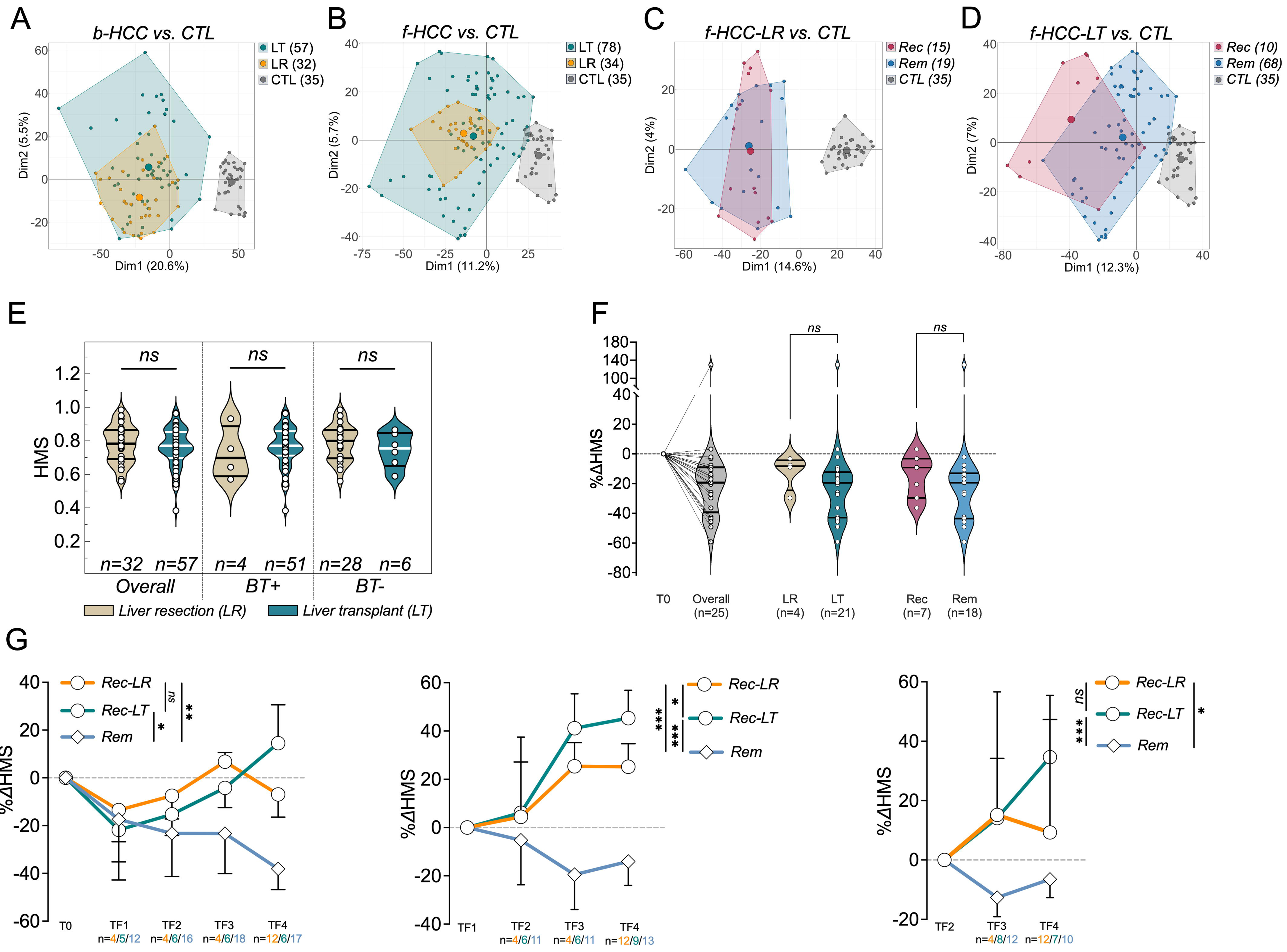
HMS comparison between liver resection (LR) and transplantation (LT) (A-D) PCA plots for baseline HCC (b-HCC, A) and follow-up HCC (f-HCC, B) samples from the patients who received liver resection (LR) or liver transplantation (LT). For f-HCC samples, whether after resection (f-HCC-LR, C) or transplantation (f-HCC-LT, D), cluster analysis was performed separately. These analyses were categorized based on the postoperative outcomes of recurrence (Rec) or remission (Rem). Healthy control samples (CTL) are included in each cluster analysis. The centers (mean points) of each cluster are represented by larger circles, number of samples were indicated next to the legend. PCA analysis was conducted based on the 4994 cancer-specific DMRs identified during HCC classifying. (E) Violin plots showing the comparison of b-HCC HMS values between liver resection (LR) and liver transplantation (LT) for all patients (Overall), as well as for those who underwent preoperative bridging therapy (BT+) or not (BT-). (F) Violin plots illustrate the overall %ΔHMS distribution for 25 patients in their first follow-up samples (FU-1, within 6 months post-surgery) normalized to T0 (Grey). These samples are grouped for comparison between patients who received resection (LR) vs. transplantation (LT) (middle), and between recurrence (Rec) vs. remission (Rem) (right). n indicates the number of cfMeDIP tests. Non-parametric Mann–Whitney U test was applied to determine whether two groups were statistically different. ns indicates no significance. (G) %ΔHMS in each group normalized to T0 (left), TF1 (middle) and TF2 (right) for recurrence and remission (Rem) patients during postoperative timeframes. Recurrence samples were sub-grouped based on the surgery type (Rec-LR, liver resection; Rec-LT, transplantation). Symbols and error bars indicating median ± IQR. Ordinary two-way ANOVA (main effects only method) was applied to determine whether there were significant divergences between two curves over time. n indicates the number of cfMeDIP tests with the corresponding color of group. ns indicates no significance. * p < 0.05, ** p < 0.01, *** p < 0.001, **** p < 0.0001. IQR, interquartile range. Source data provided as supplementary file.

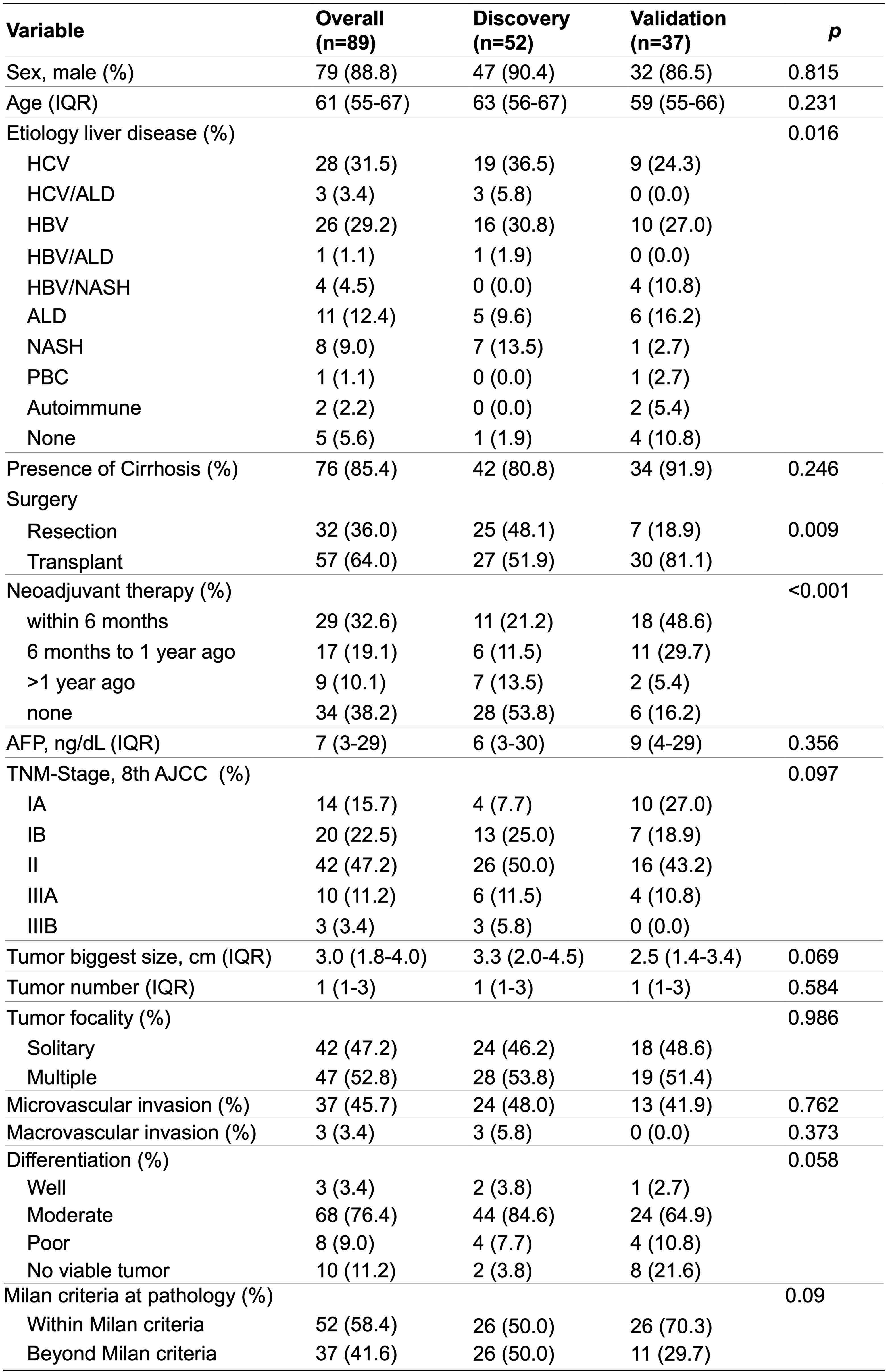

## METHODS

The overall study design is depicted in Supplementary Fig. S1.

### Patient Cohort

The study was approved by the University Health Network Research Ethics Board (REB#08-0697, REB#17-5925). All patients with liver lesions suspicious for HCC were managed in accordance with internationally accepted guidelines [23]. Between April 2014 and July 2023, patients were recruited to the study once a diagnosis of HCC was established and a decision was made to proceed with surgical resection or listing for liver transplantation [24]. Living liver donors were also recruited as healthy controls, following completion of an extensive evaluation that revealed a normal liver and absence of any systemic illness including cancer [25]. Written informed consent was provided by all study participants. Clinicopathologic data was extracted from patient charts. For liver transplant recipients, immunosuppression was based on calcineurin inhibitors (tacrolimus or cyclosporine) and corticosteroids. Corticosteroids were tapered over the first 3 months after transplantation. Recipients of living donor grafts also received basiliximab induction therapy.

### Sample collection and processing

Blood was collected in Cell-Free DNA BCT tubes (STRECK, 218997). For baseline samples from HCC patients or healthy living liver donors, blood was collected from the central venous catheter after induction of general anesthesia but before initiation of the surgical procedure. Postoperative follow-up samples were collected from HCC patients by peripheral venipuncture. Samples were processed within 2 hours of collection. Plasma supernatant was collected after 10 min of 1,900 x g centrifugation at 4°C with soft deceleration ramp then stored at −80°C until further processing. The ctDNA was isolated using QIAamp Circulating Nucleic Acid Kit (QIAGEN, 55114) followed by Qubit dsDNA High Sensitivity Assay (ThermoFisher, Q32854) quantification test and stored at −80°C.

### cfMeDIP-seq

For each sample, a total of 10 ng input ctDNA was subjected to library preparation using KAPA HyperPrep Kit (Roche Diagnostics, KK8504), customized UMI duplex adapters [26], MagMeDIP qPCR (Diagenode, C02010021) and IPure v2 kit (Diagenode, C03010015) followed by previously published 5-mC monoclonal antibody-based protocols [27, 28]. To guarantee a consistent immunoprecipitation (IP) specificity and efficiency, a single unique batch of 5mC-specific antibody (Diagenode, C15200081, Lot# RD001D, clone# 33D3, RRID: AB_2572207) was applied across all library preparations. To minimize batch effects resulting from library preparation handling, serial samples from unique patients were processed together in a single batch (maximum 24 samples per batch). The success of IP was evaluated by detecting the recovery of the spiked-in methylated and unmethylated A. thaliana DNA (Diagenode) using qPCR. Libraries that failed to pass the quality-control threshold (Fig. S2A, left) [28] were re-prepared. The number of PCR amplification cycles was determined using the Library Quant DNA Standard (NEB, E7630S). Amplified cfMeDIP libraries were purified by dual-size selection including 0.6x-followed by 1.2x-volume of AMPure XP beads (BECKMAN COULTER, A63882). Library size distribution and absence of adaptor contamination were verified by Bioanalyzer trace assay before sequencing on Illumina NovaSeq 6000 or HiSeq 2500 platform with paired-end 150-bp or 125-bp reads for ∼100 million reads per sample.

### Quality control and processing of sequencing reads

The quality and quantity of post-sequencing raw FASTQ files were examined using FastQC v0.11.5 [29] and MultiQC v1.7 [30]. Pre-processed reads were filtered using fastp v0.23.1 [31] with customized settings in paired-end mode, which included specified adapter/spacer trimming, base correction, global trimming, biased composition plus insert fragments size distribution reporting. This process was applied to a total of 238 plasma cfDNA samples (90 b-HCC, 113 f-HCC, and 35 CTL) as part of a cfMeDIP-Seq study aimed at profiling to an average depth of 101-129 (median 113) million reads. However, two prepared libraries (1 b-HCC and 1 f-HCC) were excluded due to abnormal curves identified in insert size distribution (ISD) testing results (Fig. S1, Fig. S2B, 2C).

The remaining samples, comprising 89 b-HCC, 112 f-HCC, and 35 CTL, successfully passed all cfMeDIP-seq quality control metrics and were further aligned to the hg19 reference genome using Bowtie2 v2.4.5. [32], paired reads with insert size <50-bp or above the second observed (500-bp) peak were removed synchronously [28]. SAM alignment files were converted to BAM format followed by sorting and indexing using SAMtools v1.14 [33]. Duplicated sequences from BAM files were collapsed by sambamba v0.7.0 [34] Quality control of post-processed BAM files was assessed by Qualimap v2.2 [35] as well as various metrics obtained from the R package [36] MEDIPS v 1.50.0 [37] including Saturation (MEDIPS.saturation), CpGs coverage (MEDIPS.seqCoverage) and CpG enrichment (MEDIPS.CpGenrich) analysis (Fig. S2A, right). Sample identities (b-HCC and corresponding f-HCCs) from the same individual were verified using NGSCheckMate v1.0.0 [38]. Raw counts matrix was generated from the wig (Wiggle Track Format) file exported from each BAM file using MEDIPS.exportWIG (MEDIPS) at 300-bp window size. Library and sequencing platform batch effects were reduced using sva/ComBat-seq v3.46.0 [39]. The 20% of lowest expressed counts were filtered out [17] before CPM (counts-per-million) or Trimmed Mean of the M-values (TMM) counts normalization using R packages edgeR v3.28.0 [40] and limma v3.42.0 [41]. Reads falling in ENCODE blacklist regions [42] were also excluded.

### HCC classification and subtyping using machine learning approaches

As a discovery cohort to identify HCC-specific signatures, we utilized baseline samples from 35 healthy controls and 52 HCC patients that did not have any postoperative follow-up blood collection. A stratified 10-fold cross-validation strategy was employed (Fig. S1). In each iteration, 90% of the samples from the total dataset were regarded as training data, while the remaining 10% were used as testing data to identify HCC-specific DMRs using the moderated t statistic with limma-trend method [15,16]. 300-bp size DMRs were detected by first binning the genome into 300-bp windows and then testing each bin for differential methylation between baseline HCC and healthy control groups. Model optimization was performed using three rounds of 10-fold cross-validation in the training cohort only and model performance was evaluated by computing the area under the receiver operating characteristic (ROC) curve (AUROC) using R package pROC v1.18.0 [43].

The top 300 DMRs (top 150 DMRs with gain in HCC and the top 150 DMRs with loss in HCC relative to CTL) were selected as features, and a random forest classifier (R packages caret v6.0, randomForest v4.7.1.1 and glmnet v4.1.7) [44–46] was trained based on the 9 training subsets samples to predict the probabilities of testing samples being HCC. The entire cross-validation process was repeated 100 times for a total of 1000 differential methylation analyses. For each sample, an HCC Methylation Score (HMS) was calculated as the mean probability across methylation bins associated with the HCC-specific DMRs, providing an estimate of the probabilities (ranging from 0 to 1) that a sample is classified as HCC. The HMS corresponds to the probability that the cancer specific DMRs are identified in the corresponding sample and that the patient has HCC.

To evaluate the accuracy of the HCC classifier and HMS, we utilized a validation cohort consisting of another 37 patients from whom baseline (b-HCC, n=37) and serial follow-up samples (f-HCC, n=112) had been collected (Fig. S1).

To subtype HCC, 78 baseline HCC samples (grouped based on the major liver disease categories including alcoholic liver disease (ALD), hepatitis B (HBV), hepatitis C (HCV), metabolic dysfunction-associated steatohepatitis (MASH), and no known liver disease (ND)) plus 35 healthy controls (CTL) were included in training-testing modeling according to the previously published one-vs-other strategy [16]. The combined cohort was randomly split into 100 different training-test (8:2) sets balanced for each class. Training sets were exclusively used to identify the top 300 DMRs distinguishing each class from the others. These DMRs then informed the training of a suite of ‘one versus another’ regularized, generalized linear models. The efficacy of each model was gauged by calculating the AUROC for each sample withheld in the test cohorts. An analysis was also carried out to exclude the possibility that the DMRs comprising the HCC signature were related to the presence of cirrhosis rather than cancer. This was achieved by comparing samples from patients with HBV-related HCC that arose in both cirrhotic and non-cirrhotic livers.

### Analysis of postoperative HMS trajectory and recurrence prediction

Simple moving average (SMA) analysis was conducted to compare HMS values in patients with recurrence and remission within a time-series dataset including all patients that had both baseline and follow-up samples available. The dataset was sorted by follow-up time points, and moving averages were calculated over a window size of 8.6 weeks (2 months) to reduce short-term fluctuations and highlight long-term trends. This involved averaging HMS values across each window, with the resulting averages plotted over time to visually compare trends and variations between recurrence and remission groups. Timeframes (TFs) were established to represent the significant interval differences of HMS values between recurrence and remission groups. ΔHMS was calculated as the percentage change in HMS values longitudinally before and after a reference point within the same sample with pairwise comparison strategy. Trends in *Δ*HMS were assayed for their association with postoperative recurrence and remission as confirmed by clinical methods such as imaging, AFP, and biopsy.

### Statistical analyses and illustrations

The clinicopathology statistical analysis focused on the primary outcome of RFS using Kaplan-Meier method (log-rank tests) were conducted using R software [36] and GraphPad Prism v9.5. Survival times were calculated from the date of the surgery (either liver resection or transplant) to either the date of death or the last follow-up, with the cutoff for the last follow-up being October 25, 2023. The analysis included both categorical and continuous variables. Categorical variables were presented as numbers and percentages, while continuous variables were expressed as median ± IQR. For comparative analysis, the Chi-square test was used for categorical variables and the Mann-Whitney U test for continuous variables. To evaluate the impact of HMS > 0.9 on tumor recurrence risk, an a priori multivariable Cox proportional-hazard regression was employed, with results presented as HR and 95%CI. Model accuracy was assessed using ROC analysis under the logistic model, comparing the AUC.

Other statistical analysis, including descriptive statistics (Quartiles analysis), normality and lognormality (D’Agostino & Pearson method) test, outlier identification were performed and visualized using software GraphPad Prism 9.5. p < 0.05 was considered statistically significant. Scatter plot, box-whiskers plot results were represented as mean ± SD for normally distributed variables or median ± IQR for non-normally distributed variables. Non-parametric Mann– Whitney U test was applied to determine whether two groups were statistically different for non-parametric variables. Principal Component Analysis (PCA) and visualization were conducted using R packages Factoextra v1.0.7 [47] and FactoMineR v2.8 [48]. Graphs were illustrated using GraphPad Prism and R package ggplot2 v3.4.2 [49].

## ABBREVIATIONS

AFP: α-fetoprotein
AIC: akaike information criterion
ALD: alcoholic liver disease
AUROC: area under the ROC curve
BAM: binary alignment map
b-HCC: baseline HCC
BT: bridging therapy
cfDNA: cell-free DNA
cfMeDIP-Seq: cell-free methylated DNA immunoprecipitation and high-throughput sequencing
CI: confidence interval
CPM: counts per million
CT: computed tomography
ctDNA: circulating tumor DNA
CTL: healthy control
DMR: differentially methylated region
f-HCC: follow-up HCC
f-HCC-Rec: follow-up samples from patients after being diagnosed with HCC recurrence
f-HCC-Rem: follow-up samples from patients during their non-recurrence clinic follow-up recording time
HBV: hepatitis B virus
HCC: hepatocellular carcinoma
HCV: hepatitis C virus
HMS: HCC methylation score
HR: hazard ratio
IQR: interquartile range
IRE: irreversible electroporation
ISD: insert size distribution
LR: liver resection
LT: liver transplantation
LTD: largest tumor diameter
MASH: metabolic dysfunction-associated steatohepatitis
MiVI: microvascular invasion
MRI: magnetic resonance imaging
MWA: microwave ablation
NA: not applicable
ND: no known liver disease
NT: non-treatment
PBC: primary biliary cholangitis
PCA: principal component analysis
PEI: percutaneous ethanol injection
QC: quality control
qPCR: quantitative polymerase chain reaction
RFA: radiofrequency ablation
RFS: recurrence-free survival
ROC: receiver operating characteristic
SAM: sequence alignment map
SBRT: stereotactic body radiation therapy
SEM: standard error of the mean
SMA: simple moving average
TACE: transcatheter arterial chemoembolization
TF: timeframe
TMM: trimmed mean of M-values

## DATA AND CODE AVAILABILITY

Deidentified data are provided in the Supplemental Tables. Raw sequencing data for all 203 HCC samples and 23 healthy controls are deposited at the European Genome-phenome Archive (EGA) under the accession number EGAS50000000450. Raw sequencing files for another 12 healthy normal controls can be requested at https://www.ontariohealthstudy.ca/for-researchers/data-access-formsand-templates. Processed intermediate data and objects used to replicate the results in this study are available at https://doi.org/10.5281/zenodo.11251606. Code scripts and markdown instruction to reproduce the figures and analyses are available at https://github.com/pughlab/HCC_cfMeDIP.

## ACKNOWLEDGMENTS

This study was conducted with support from the Canadian Institutes of Health Research (CIHR Project grant #152982, T.J.P., S.P.C., and A.G.) and the UHN Lillian MacKenzie Foundation (A.G.). T.J.P. holds the Canada Research Chair in Translational Genomics and is supported by a Senior Investigator Award from the Ontario Institute for Cancer Research and the Gattuso-Slaight Personalized Cancer Medicine Fund. We thank members of the Princess Margaret MeDIP-seq working group for helpful discussion around analysis methods, particularly Emma Bell, Samantha Wilson, Justin Burgener, Eric Zhao, Nick Cheng, Ming Han, and Sasha Main. We thank the staff of the Princess Margaret Genomics Centre (www.pmgenomics.ca), UHN Bioinformatics and HPC Core (https://bhpc.uhnresearch.ca), and the Ontario Institute for Cancer Research Genomics Program (genomics.oicr.on.ca). These programs were enabled through funding provided by the Government of Ontario and the Princess Margaret Cancer Foundation. Additional infrastructure support from the Canada Foundation for Innovation, Leaders Opportunity Fund (CFI #32383 and #38401); the Ontario Ministry of Research and Innovation, Ontario Research Fund Small Infrastructure Program; and the Ontario Institute for Cancer Research.

